# Analysis of genetic overlap between inborn errors of immunity and neurodevelopmental disorders

**DOI:** 10.1101/2025.03.01.25323148

**Authors:** Ines Serra, Mick de Koning, Peter J. van der Spek, Virgil A.S.H. Dalm, Aleksandra Badura

## Abstract

Inborn errors of immunity (IEI), formerly known as primary immune deficiencies (PID), are a group of genetic disorders that affect the immune system, leading to increased susceptibility to infections, autoimmunity, allergy, and cancer. So far, 449 IEI-causing genes have been identified with more likely to be discovered with the rapid adoption of whole genome sequencing in clinical practice. Patients with IEI often present with neurological symptoms such as cognitive impairments, neurodevelopmental delay and even seizures. These clinical features could be indicative of an increased risk of neurodevelopmental disorders (NDDs) in IEI patient population. However, to date, no exhaustive study has been done on the genetic overlap between NDDs and IEIs. Using publicly available NDD and IEI variant databases, gene ontology analysis, machine learning, and protein-network clustering analysis, we found that one-third of IEI-causing genes were also linked to NDDs. These genes were primarily involved in immune development and DNA repair pathways. In contrast, genes causing exclusively IEIs were enriched in immune response functions. Functional connectivity analysis revealed that NDD-risk genes integrated immune-related networks, including those involved in DNA repair, highlighting immune-NDD interactions. Altogether, this work demonstrates a molecular and protein-network level overlap between NDD and IEI-causing genes. Our analysis strongly suggests that NDD phenotypes in IEI patients could be underreported in NDD-related databases.

## Introduction

Inborn errors of immunity (IEIs) are a group of heterogeneous genetic disorders, mostly caused by monogenic variants, that affect the capacity of the immune system to recognize, respond to and resolve immunological threats (Shields and Patel 2017). Over the past 40 years, the identification of genetic variants causing IEI has been facilitated with the advent of next-generation sequencing and with the increased accessibility to these technologies in clinical settings (Tangye et al. 2022; Fusaro et al. 2021). As of January 2025, 485 variants known to cause IEI have been officially recognized (Tangye et al. 2022). Notably, rates of genetic testing are highly variable worldwide and its widespread use is still infrequent in many settings, particularly in low to middle income countries (Aghamohammadi et al. 2021). Therefore, for a number of IEIs, including those where a causative gene is yet to be identified, diagnoses still rely heavily on clinical presentation and laboratory workup (Anderson et al. 2022).

Despite increasing availability of genetic sequencing, the identification of IEIs often comprehends a considerable delay between first clinical presentation and actual diagnosis (M. G. Lawrence et al. 2024). For example, a median diagnostic delay of 4 years was reported amongst 2700 patients with common variable immune deficiency (CVID), the most prevalent IEI, thought to affect 1:25.000 people (Odnoletkova et al. 2018). Diagnostic delay is therefore a major contributor for late therapy initiation, even though 2/3 of IEI cases are still diagnosed during pediatric ages (Thalhammer et al. 2021; Quinn et al. 2022). Importantly, the increasing number of patients diagnosed with IEI reveals that this group of disorders is far more common than previously thought, with a predicted prevalence between 1:1.000 and 1:5.000 individuals (Abolhassani et al. 2022; Akalu and Bogunovic 2024). Thus, although individually rare, IEIs significantly contribute to societal and economic health-related burden.

Despite phenotypic variability, patients with IEIs generally experience an increased risk of infections and immune dysregulation, often in the form of autoimmunity (Thalhammer et al. 2021). Additionally, symptoms including enteropathy, failure to thrive, atopy and increased risk for malignancies are also commonly seen in these patients (Mohammadi et al. 2023; Rasouli et al. 2023). For a number of IEIs, neurodevelopmental and cognitive deficits (from here on referred to as neurodevelopmental deficits, NDDs) have also been reported (Kurup et al. 2024; Manusama et al. 2022). While this symptomatology has long been recognized as a component of disease presentation for some groups of IEI patients, as is the case of those with DiGeorge syndrome (Nissan et al. 2021), a growing number of cohort studies examining other IEIs has reported neurodevelopmental and cognitive phenotypes in these patients at higher prevalences than those found in reference populations (Zorn et al. 2022; Elkaim et al. 2016; Coulter et al. 2017; Cagdas et al. 2018; Kerr et al. 2010).

Chronic conditions such as IEIs, which are accompanied by frequent hospitalizations, have been shown to contribute to delays in development, particularly with regards to the presence of disruptive behaviours, learning capacity, attention, and social participation (Fardell et al. 2023). Furthermore, in adult patients with IEI, “brain fog” is a commonly reported neurocognitive impairment that limits attention and overall cognitive performance (Sowers, Gayda-Chelder, and Galantino 2020). Thus, the presence of IEIs alone, particularly those starting at young ages, increases the developmental vulnerability of patients in several cognitive domains. However, for a number of behavioural and cognitive aspects, the negative impact of IEIs can still be found when patients’ chronic condition status is accounted for or even after curative treatments, such as hematopoietic stem cell transplantation, have been implemented (Titman et al. 2008; Kerr et al. 2010; Nicholson et al. 2022). Together, this suggests that neurodevelopmental and cognitive symptomatology in patients with IEI is likely not simply a secondary consequence of hospitalization and chronic disease burden but specific consequence of genetic variations that affect brain function.

This interaction between immune and nervous systems can also be seen through the perspective of NDDs, where people often present with immune dysregulation. Children with autism spectrum disorder (ASD), one of the most common NDDs, appear to be more susceptible to infections, particularly during the early years of life (Hall et al. 2023; Sabourin et al. 2019), and are also more likely to exhibit food, skin and respiratory allergies (Xu et al. 2018). Furthermore, transcriptomic alterations in immune genes have also been reported in people with ASD. In a study using cortical samples from individuals with autism, a significant upregulation of genes involved in immune and inflammatory response was found (Voineagu et al. 2011), while data from single nucleus RNA sequencing shows that brain tissue from people with ASD have increased numbers of reactive microglia, and upregulated neural-immune receptors as well as inflammatory response effectors (Wamsley et al. 2024). Additionally, GWAS meta-analyses have demonstrated associations between autistic traits and genes involved in the regulation of immune response (Arenella et al. 2021), as well as up to 30 genomic loci where distinct NDDs exhibit significant associations with immune disorders (Xiu et al. 2024). Altogether, these studies suggest that a possible mechanism driving the development of NDDs in patients with IEIs could be the presence of genetic variants that affect not just the immune system but also brain function. However, the extent to which IEI-causing genes are implicated in NDD symptomatology is currently unknown, preventing the identification of common pathways between IEIs that could potentially predict the risk of NDD development.

To gain a deeper understanding of putative shared mechanisms that could underpin the coexistence of immune and nervous systems phenotypes, we analyzed gene expression of NDD-risk and IEI-causing genes. Specifically, using a combination of available sequencing databases, gene ontology analysis, machine learning approaches and protein-network reconstruction, we investigated whether genes previously associated with NDD, NDD and IEI, or only IEI phenotypes presented with specific expression patterns or activated distinct molecular networks. We found that a third of IEI-causing genes had been associated with NDD symptomatology (i.e., *shared* genes) and were often associated with nuclear-heavy functions namely immune system development and DNA-repair. Contrastingly, genes associated with only IEIs were primarily involved in immune system response. Furthermore, although cellular expression patterns were insufficient to classify gene groups, high-risk syndromic autism genes could be separated from IEI-causing genes, while shared genes were often misclassified as IEI, indicating common cellular expression patterns. Finally, reconstruction of functional connectivity uncovered the involvement of NDD-risk proteins in biological processes commonly associated with immune function.

Altogether, our results add to a growing body of evidence supporting vast interactions between immune and nervous system genetics and provide novel insights into IEI subgroups that could potentially benefit from a systematic NDD evaluation during their assessment and treatment plan.

## Results

### IEI and NDD risk genes show a significant overlap

To explore the putative commonalities between IEI and NDD risk genes, we analyzed two distinct databases. First, we used the 2022 update on IEI classification from the International Union of Immunological Societies (IUIS) Expert Committee to collect all current genes recognized as causative of IEI (Tangye et al. 2022). After removing duplicates (for example, the *TYK2* gene can be involved in two distinct disorders, TYK2 deficiency and P1104A TYK2 homozygosity), we obtained a list of 449 genes (*“IEI genes”*). We then surveyed the Human Gene Mutation Database (HGMD) for genes associated with “neurodevelopmental disorder”. After the elimination of duplicates and gene nomenclatures not recognized by Human Genome Organization (HUGO) gene nomenclature committee, a total of 6003 genes were retained for further analysis (*“NDD genes”*) (**Figure 1A**). Next, we calculated the overlap between the two lists to identify the risk genes that were shared across the two databases (**Figure 1B**). We found a statistically significant overlap (p < 0.0001) between the NDD and IEI risk gene groups (*“shared genes”*) with a representation factor of 3.4, indicating a larger overlap than theoretically expected for two randomly chosen gene lists of this size (Hirbo et al. 2015). In total, approximately 33% of IEI-causing genes were shared with NDD-associated genes, while 2.5 % of the NDD-associated genes were found among the IEI-causing genes.

**Figure 1:**
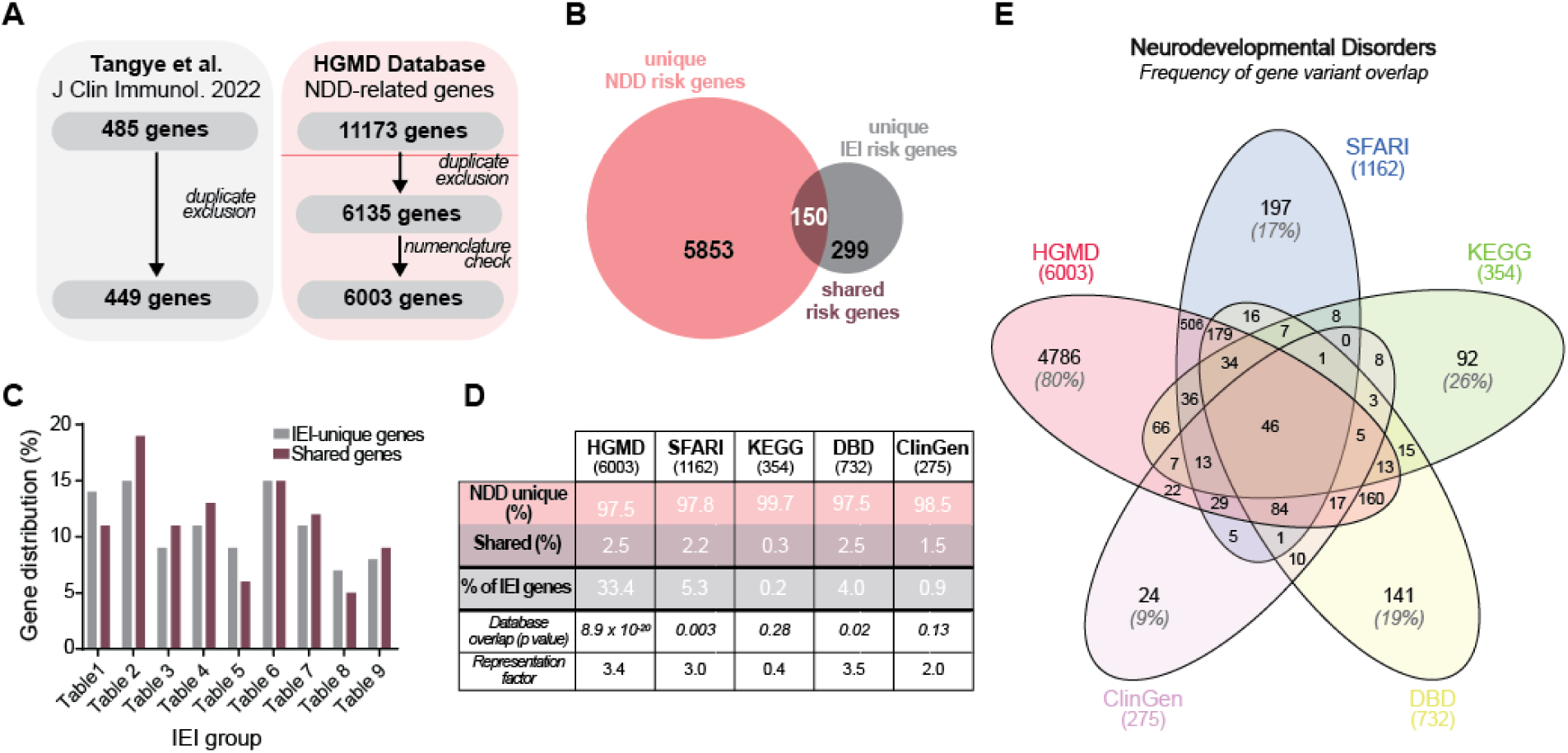
Overlap between genetic variants leading to NDD and IEI development. **A)** Gene selection schematics. **B)** Calculated overlap between the HGMD and IEI gene lists. **C)** Distribution (in percentage) of the total IEI-risk genes and the identified shared genes across the IUIS phenotype classification tables. Note that table 10 was omitted from the analyses as it combines somatic mutations with auto-antibody production phenotypes. **D)** Percentage of identified genes that are unique to each curated database (pink) and that overlap with the IEI gene list (purple); percentage of IEI risk-genes identified in each database. **E)** Venn diagram demonstrating the genetic overlap between databases (percentages of unique, non-overlapping, genes are shown in grey).

The IUIS classifies IEI into 10 distinct categories organized into tables that aim to group IEIs with similar phenotypes (Bousfiha et al. 2022) (**Supplementary Table 1**). To understand whether *shared* genes were associated with a specific IEI category, we calculated the observed and expected distribution of these genes within IEI groups. Genetic variants categorized as “*Combined immunodeficiencies with associated or syndromic features*” (Table 2 of the IUIS classification) encompasses the highest number of identified *shared* genes. However, we found that the observed distribution of *shared* genes within each IEI table was not statistically different from the distribution of all IEI genes across tables (*X^2^*(8) = 6.644. p = 0.57) (**Figure 1C**). These results suggest that, when using the IUIS IEI classification, no single category presents with an enrichment of the identified *shared* genes.

The pool of NDD risk genes obtained from the HGMD database comprises an extensive list of genes associated with wide-ranging neurodevelopmental disorder phenotypes, including but not restricted to: autism spectrum disorder (ASD), intellectual disability, speech and motor delay. Therefore, to better understand the genetic overlap between the IEI and NDD genes we included in our analysis other clinically relevant databases, a number of which had been manually curated towards specific neurodevelopmental disorders. Specifically, we investigated the presence of IEI genes within the KEGG Disease (*“neurodevelopmental disorder”* classification), ClinGen (Intellectual Disability and Autism Expert Panel), Developmental Brain Disorder Gene Database (DBD) and the SFARI database (focused on the curation of risk genes associated with ASD). On average, each of these databases presented with a percentage of unique genes (i.e. not present in the other databases) of approximately 18%, with the remaining genes overlapping between databases to varying degrees (**Figure 1E**). Similar to what we identified with the HGMD database, the overlap of IEI genes with both the SFARI and DBD databases was also statistically significant (**Figure 1D**), with a representation factor > 1 indicating an overlap larger than expected given a random selection of two groups of human genes. The overlap with the ClinGen and KEGG databases did not reach statistical significance (**Figure 1D**). Altogether, this data suggests that a subgroup of genetic variants known to cause IEI are also associated with heterogeneous clinical phenotypes typically associated with NDDs.

### Immune cell development and DNA repair processes distinguish shared from IEI-unique genes

Next, we used gene enrichment analysis to investigate the molecular pathways and biological functions in which the previously identified gene groups (*NDD-unique*, *shared* and *IEI-unique*) were involved. In order to summarize relevant and enriched biological processes with minimal redundancy (Liberzon et al. 2015), we first used the “*Hallmark gene sets*” subcollection from the Human Molecular Signatures Database (MSigDB), followed by enrichment analysis using the Gene Ontology (GO) Consortium knowledgebase to retrieve statistically significant annotations on biological processes and cellular components associated with the genes of interest (Ashburner et al. 2000; Gene Ontology Consortium et al. 2023).

We found that the *NDD-unique* gene list was enriched for a number of signaling pathways involved in overall organism development. The highest enrichment values were found for the Hedgehog and Notch pathways, essential for stem cell maintenance and cell identity specification in the central nervous system, respectively (Briscoe and Thérond 2013; Zhou et al. 2022; Ferent et al. 2014) (**Figure 2A, left**). Accordingly, *NDD-unique* genes appeared enriched in biological processes integrating neuronal development and the biogenesis of polarized substructures, essential for neuronal development (**Figure 2B, left**). This enrichment towards brain-related processes was also captured by the Cell Components GO, in which a high number of genes was associated with key neuronal structures (**Figure 2C, left**).

**Figure 2:**
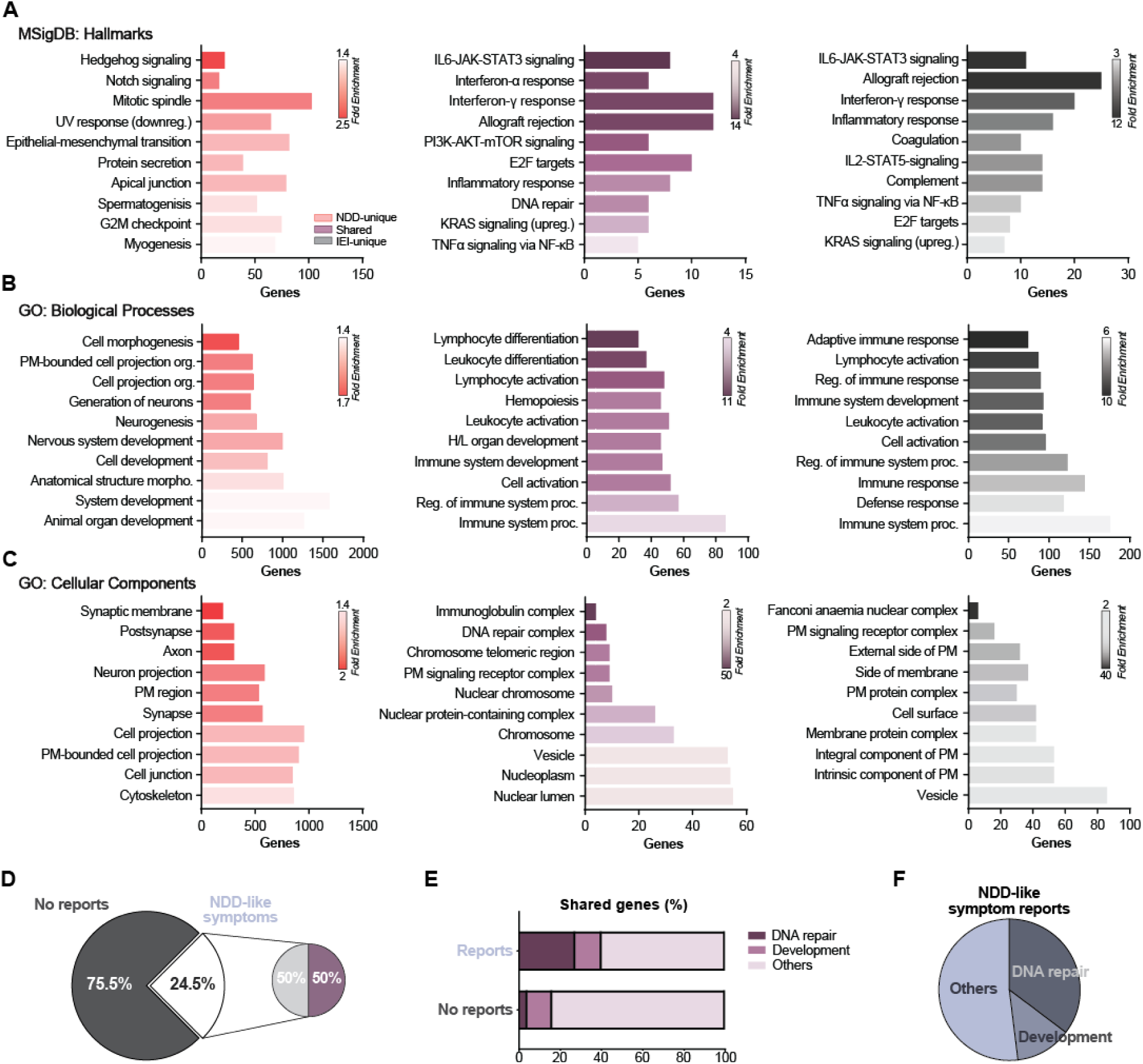
Gene Ontology (GO) analysis and NDD-like symptom distribution distribution. Bar charts, ranked by fold enrichment over the number of identified genes, showing the top 10 GO terms for **A)** the Human Molecular Signatures Database Hallmark Gene set, **B)** biological processes and **C)** cellular components, for all identified NDD-unique genes (left), shared genes (centre) and IEI-unique genes (right). **D)** Percentage of genes with NDD-like symptomatology reported in the IUIS classification tables. **E)** Percentage of identified shared genes with DNA repair or development functions described in the IUIS classification tables. **F)** Distribution of DNA repair or development functions in genes associated with NDD-like symptoms in the IUIS classification tables.

Both *IEI-unique* and *shared* genes were involved in a number of signaling pathways within the immunological response. While both gene groups presented with similar classes of enriched molecular pathways (**Figure 2A, centre & right**), *IEI-unique* genes were implicated in a number of processes related to immune response itself, with terms representative of the activation and effector phases of immune response identified in the Biological Processes GO (**Figure 2B, right**). Most enriched cellular components for these *IEI-unique* genes were located at the level of the plasma membrane, representative of the enrichment of highly specialized transmembrane proteins and cell surface receptors characteristic of most cell populations comprising the immune system, particularly at the level of the immunological synapse (Céspedes et al. 2021) (**Figure 2C, right**). Contrastingly, most *shared* genes did not appear to have a significant involvement in these reactive processes. Most of the identified biological processes for the *shared* genes were enriched in terms related to the development and differentiation of immune system cells (**Figure 2B, centre**) (Dorshkind and Crooks 2023; Katharina Simon, Hollander, and McMichael 2015). These distinct biological functions were also captured in the identified molecular pathways, with the PI3K-AKT-mTOR pathway identified as enriched in the *shared* but not the *IEI-unique* group (**Figure 2A, centre & right**). Notably, this pathway is not just crucial for cell division and fate determination in the immune system, but also essential for brain development, with mutations along this pathway leading to a number of neurodevelopmental disorders termed “mTORopathies” (Crino 2020; Bockaert and Marin 2015; Jones and Pearce 2017).

*Shared genes* were identified as being mostly involved with nuclear-related structures (**Figure 2C, centre**). This was consistent with the DNA-related pathways identified (**Figure 2A, centre**), and likely reflects DNA repair and DNA modification processes, such as V(D)J recombination, essential for the formation of diverse repertoires of immunoglobulins and surface receptors in developing B and T cells (Rivera-Munoz et al. 2007). Of note, mutations in DNA damage response genes have also been associated with pediatric acute-onset neuropsychiatric syndrome (PANS) (Trifiletti et al. 2022; Cunningham et al. 2024) Because DNA repair is also associated with genome instability which, as mTOR pathway dysfunction, has been implicated in NDD development, we returned to the IEI classification database by Tangye and colleagues and reviewed the described phenotype notes (“*Associated features*”) for the presence of NDD-like phenotype reports. In total, we identified 110 genes that were accompanied by descriptions of NDD-like symptoms (**Figure 2D**), representing 24.5% of all IEI genes. Of these, half represented genes we had previously classified as *shared,* while the remaining were classified as *IEI-unique*. To exclude HGMD database bias, we verified whether additional genes could be identified in the other selected databases (**Figure 1D**). Only 4 of these genes were found, namely *ADA* and *TBX1* present in the SFARI database, *TRNT1* in the DBD database, and *DKC1* in the ClinGen Database, suggesting that NDD phenotypes from these primarily immune disorders could be underreported in NDD-related databases.

Within the *shared* group, reports of NDD-like symptoms associated with genes involved in DNA repair or immune system development accounted for 40% of the entries (**Figure 2E**). Conversely, DNA repair and immune system development functions were present in approximately half of the genes for which NDD-like symptoms had been reported.

Altogether, this suggests that IEI-causing genetic variants that affect these molecular systems could potentially lead to a higher risk of development of NDD-like symptomatology.

### Genes of interest present with a progressive expression in brain and immunological human tissue

To better understand the expression patterns of IEI-causing genes and that of those associated with NDD phenotypes, we investigated distribution of the identified gene products in human tissue. To do this, we used the Human Protein Atlas (HPA) database that contained both whole tissue and single-cell RNA sequencing datasets spanning distinct systems from the human body. Specifically, we evaluated protein-coding risk-gene expression in brain-, blood, tissue- and single cell-derived datasets (Figure 3; Supplementary Figure 1 A-B).

**Figure 3:**
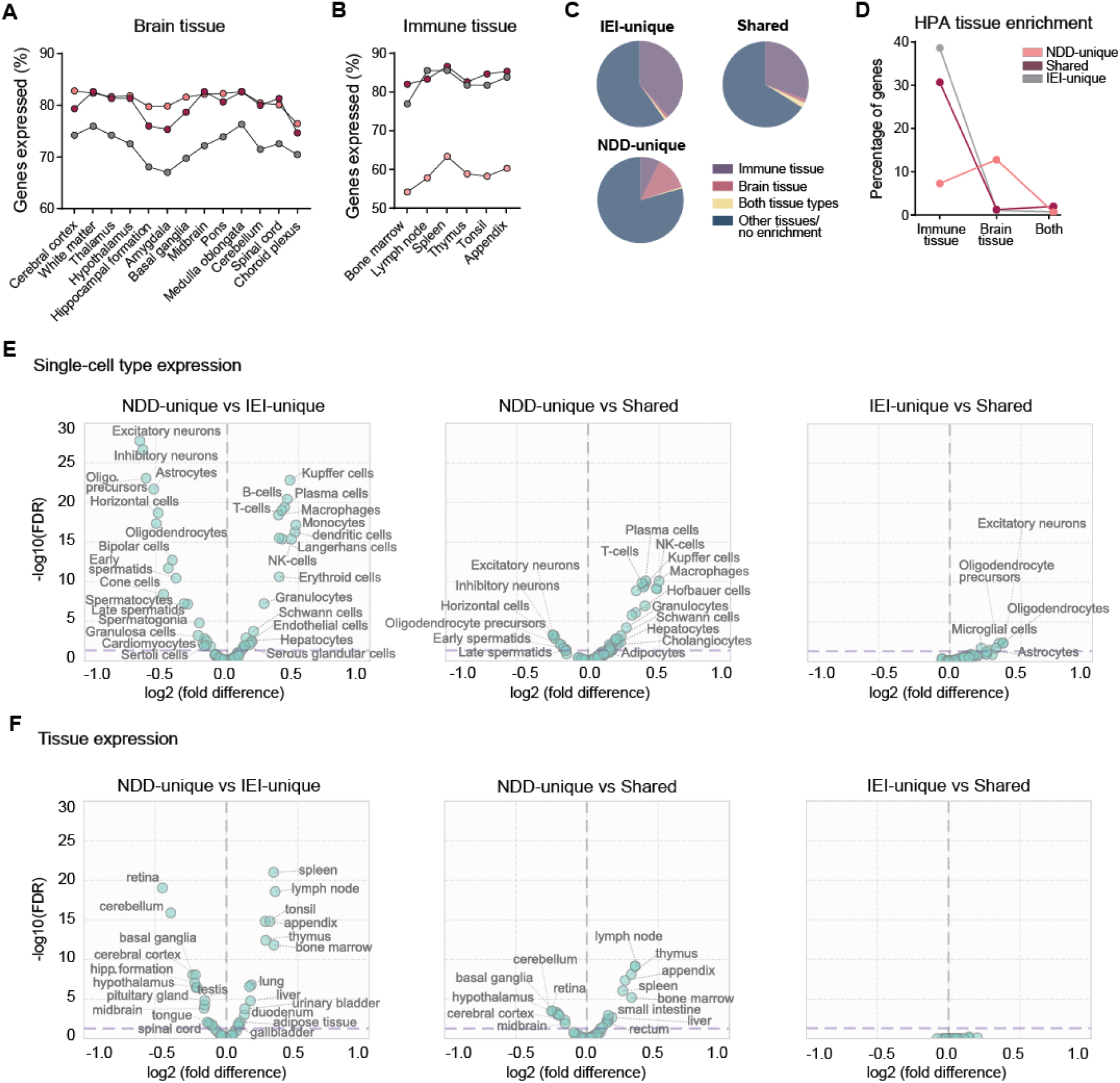
Comparative gene expression patterns across gene groups. **A**, **B)** pooled percentage of genes expressed in brain and immune tissue within the NDD-unique, shared and IEI-unique gene groups. **C, D)** HPA tissue enrichment classification and distribution across the three gene groups of interest, within the brain and immune tissue. **E, F)** Paired comparison volcano plots with gene expression fold-changes across the three gene groups (FDR < 0.05).

After identifying protein-coding genes of interest, we first investigated whether protein products were expressed in brain or immune tissues. We identified expression levels for 264 *IEI-unique* genes, most of these expressed both in brain and immune tissue (**Figure 3A-B**). Additionally, a total of 102 were enriched in lymphoid and/or bone marrow tissue and 5 genes were enriched in the brain, specifically *ITPKB*, *FCHO1* (also enriched in lymphoid tissue), *MAP1LC3B2*, *SNX10*, and *TMC6* (also enriched in lymphoid tissue). *ITPKB* was also enriched in the choroid plexus. We then looked at the site of expression of *shared* genes (**Figure 3A-B**). All 150 shared genes were identified in the HPA database and, of these, 46 were enriched in lymphoid and/or bone marrow tissue while 5 were enriched in the brain. These were *RASGRP1* (also enriched in lymphoid tissue), *CHD7*, *CARMIL2* (also enriched in lymphoid tissue), *BCL11B* (also enriched in lymphoid tissue) and *SLX4*. Finally, we analysed the tissue distribution of the 5798 *NDD-unique* gene products identified on the HPA expression database. In this group, 425 gene products were enriched in lymphoid and/or bone marrow tissue, 744 in brain tissue and 43 genes were enriched in both tissues (**Figure 3C**). Considering the distribution of enriched genes within each gene group, the highest percentage of immune tissue enrichment was found within the *IEI-unique* group, while *NDD-unique* genes presented with the highest percentage of genes enriched in brain tissue. *Shared* genes, as expected, exhibited the largest percentage of genes that were enriched both in immune and brain tissue (2% for *shared* genes vs 0.8% for the *IEI-unique* and 0.7% for the *NDD-unique* gene groups) (**Figure 3D**).

To gain a deeper understanding of how these expression patterns varied across the three gene groups, we performed comparative protein expression analysis on the HPA datasets. Within the cell-specific analysis, we found that *NDD-unique* genes presented with increased expression in CNS-related cells, such as neurons, astrocytes and horizontal cells, when compared to *IEI-unique* genes (**Figure 3E, left**). The average gene expression of *NDD-unique* genes was reduced within immune cells, including specialized macrophages, B and T cells, compared to *IEI-unique* genes. Similar expression patterns were found when comparing the cellular expression of *NDD-unique* genes with that of *shared* genes, albeit at a lower magnitude (**Figure 3E, centre**). When *IEI-unique* and *shared* genes were compared, an increased expression of *shared* genes within neuronal subtypes and glia distinguished these two genetic populations (**Figure 3E, right**).

Next, we analysed the HPA tissue expression dataset. In accordance with the single-cell dataset, we found that *NDD-unique* genes had their highest differential expression within CNS structures and lowest in lymphoid organs, when compared with *IEI-unique* genes (**Figure 3F, left**). Similar differences were also found when we compared the expression of *NDD-unique* genes with that of *shared* genes (**Figure 3F, centre**). However, contrasting with the single-cell data, we no longer detected significant differences in expression patterns between the *IEI-unique* and *shared* genes groups (**Figure 3F, right**). Lastly, blood- and brain-specific HPA datasets were also analysed. Here, most of the significant differential expression changes were found between *NDD-unique* and *IEI-unique* genes, while no variables differentiated the *IEI-unique* from the *shared* gene groups (**Supplementary figure 2**). Altogether, this data suggests that *shared* genes present with bulk cellular and organ gene expression patterns that combine features from the profiles of expression of the NDD and the IEI gene groups.

### IEI-unique, but not shared group genes, are predicted by classification analysis

Given the differences of gene expression patterns described previously, we explored whether single-cell expression values could be used to predict group allocation of *NDD-unique*, *shared* and *IEI-unique* genes. To investigate this, we first performed dimensionality reduction in the HPA single-cell dataset using the Uniform Manifold Approximation and Projection (UMAP) algorithm, which enabled the visualization of potential co-localization patterns between the three groups of genes (**Figure 4A**). This analysis revealed a large overlap in expression profiles between *NDD-unique*, *shared* and *IEI-unique* genes, suggesting considerable heterogeneity in cell type-specific expression patterns across all groups, which prevented further accurate data classification.

**Figure 4:**
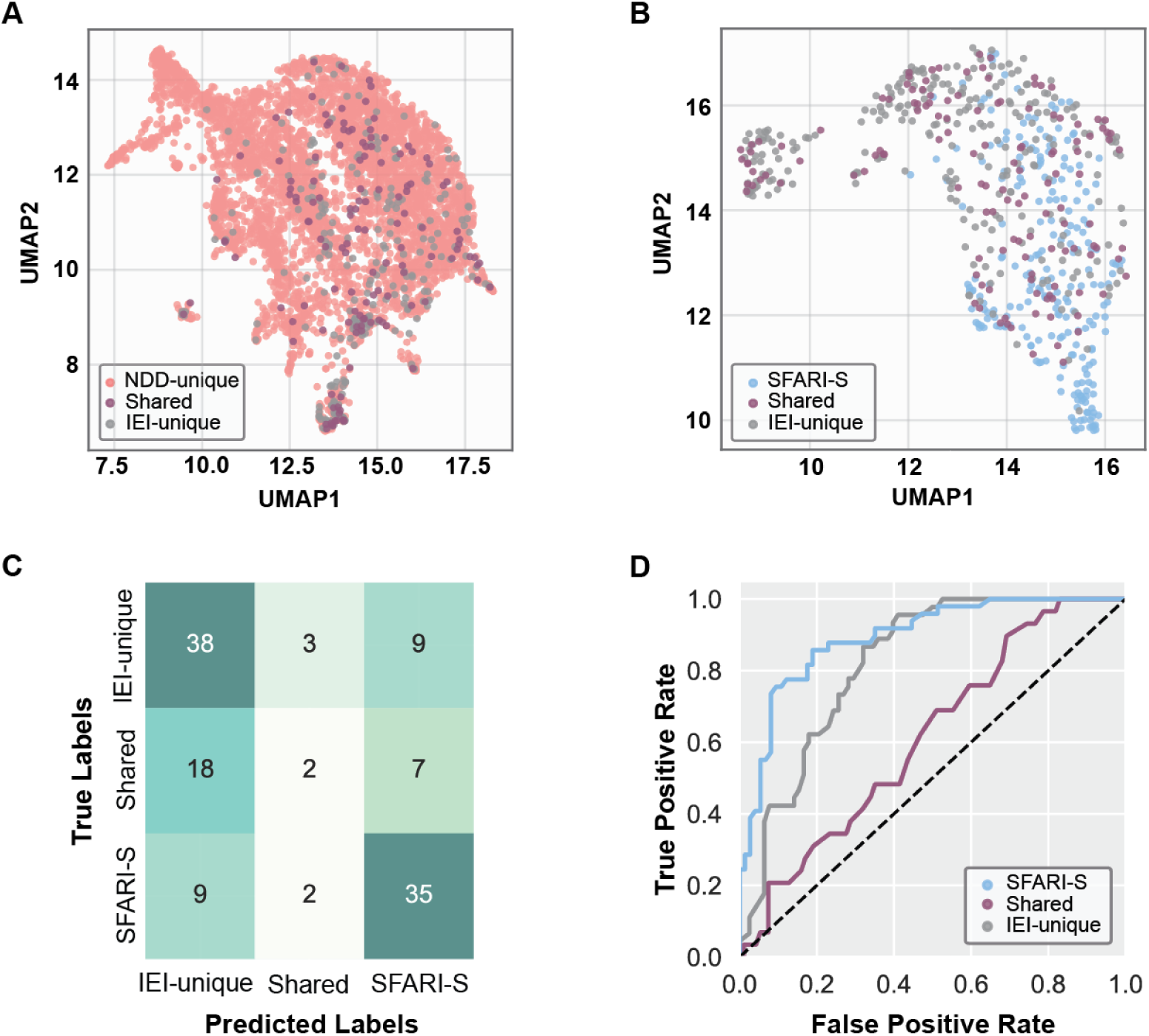
Expression Profile and Classification Analysis of Disease-Associated Genes. **A)** UMAP visualization demonstrating overlapping expression patterns between NDD-unique, shared and IEI-unique genes across cell types. **B)** Comparing SFARI-S with IEI-unique and shared genes through UMAP visualization, reveals both overlapping and distinct expression areas between groups. **C)** Random forest classifier performance metrics showing classification of testing data. SFARI-S genes (Precision: 0.69; F1: 0.72) and IEI-unique genes (Precision: 0.58; F1: 0.66) showed higher prediction accuracy compared to shared genes (Precision: 0.29; F1: 0.12). **D)** One-vs-rest multiclass receiver operating characteristic yielded AUC values of 0.89 (SFARI-S), 0.82 (IEI-unique), and 0.61 (shared). Dotted line indicates change level.

One of the reasons for this broad heterogeneity could be the fact that the HGMD database, from which *NDD-unique* genes were derived, included phenotype descriptions that cover a vast array of distinct NDDs. Thus, we next investigated whether classification could be improved using a more restricted source of NDD-risk genes. Therefore, we focused on the analysis of only high-risk autism genes, by performing dimensionality reduction analysis of syndromic-labeled genes from the SFARI database (i.e. *SFARI-S* genes). Comparison of the previously identified *IEI-unique* and *shared* genes with *SFARI-S* genes still showed variable expression patterns. However, there were areas of non-overlapping projections, indicating differential expression profiles between these groups of genes (**Figure 4B**). To understand if these potentially unique expression profiles could predict group identity, we used a random forest classifier approach. This model showed high performance in identifying *SFARI-S* (Precision: 0.69; F1-score: 0.72; Recall: 0.76), while *IEI-unique* genes exhibited moderate classification accuracy (Precision: 0.58; F1-score: 0.66; Recall: 0.76). However, *shared* genes were frequently misclassified as IEI-unique genes (Precision: 0.29; F1-score: 0.12; Recall: 0.12) (**Figure 4C**), suggesting that despite having some expression patterns in common with *NDD-unique* genes (**Figure 3**), *shared* genes are more similar to *IEI-unique* genes. Multiclass Receiver Operating Characteristic analysis further supported these findings, with area under the curve (AUC) values of 0.89, 0.82, and 0.61 for *SFARI-S*, *IEI-unique*, and *shared* genes respectively, corroborating the higher predictive value of the model on *SFARI-S* and *IEI-unique* genes compared to *shared* genes (**Figure 4D**).

### Cluster analysis reveals direct protein-protein interactions between the products of the three gene groups

To better understand putative interactions between the three groups of genes, we performed clustering analysis of a constructed protein interaction network derived from known gene-disease associations for the *NDD-unique*, *IEI-unique* and *shared* groups. We identified multiple large, highly interconnected clusters containing proteins from all three gene groups, likely corresponding to joint molecular functions (**Figure 5**). The largest cluster identified included several gene associations across the three gene groups, and showed a significant enrichment for proteins involved in biological processes related to DNA repair mechanisms. The second largest cluster demonstrated significant enrichment for immune response-related processes, including proteins involved in NF-κB signalling and T-cell proliferation. These latter processes were more commonly associated with immune related diseases, albeit showing strong interactions with NDD-unique proteins as well (**Figure 5A**). Among the remaining clusters involving more than one gene group, we observed enrichment for processes involved in cell development and migration (e.g. actin-related) and signaling events primarily involving tyrosine kinases and transcription. In contrast, clusters containing only NDD-unique proteins, without significant NDD-IEI or NDD-shared contributions, included predominantly processes associated with basic cellular functionally. This included proteins involved in synapse formation and function, as well as proteins responsible for histone modifications, ribosomal activity and Wnt signaling (**Figure 5B**). The Wnt pathway is particularly crucial for normal brain formation, and its dysregulation leads to neurological disorders and cancers. Altogether, this network analysis demonstrates the existence of complex molecular interactions between *NDD-unique* and IEI-causing genes, suggesting that network unbalance occurring in the presence of pathological genetic variants could give rise to the presence of shared phenotypes in patients with IEIs.

**Figure 5:**
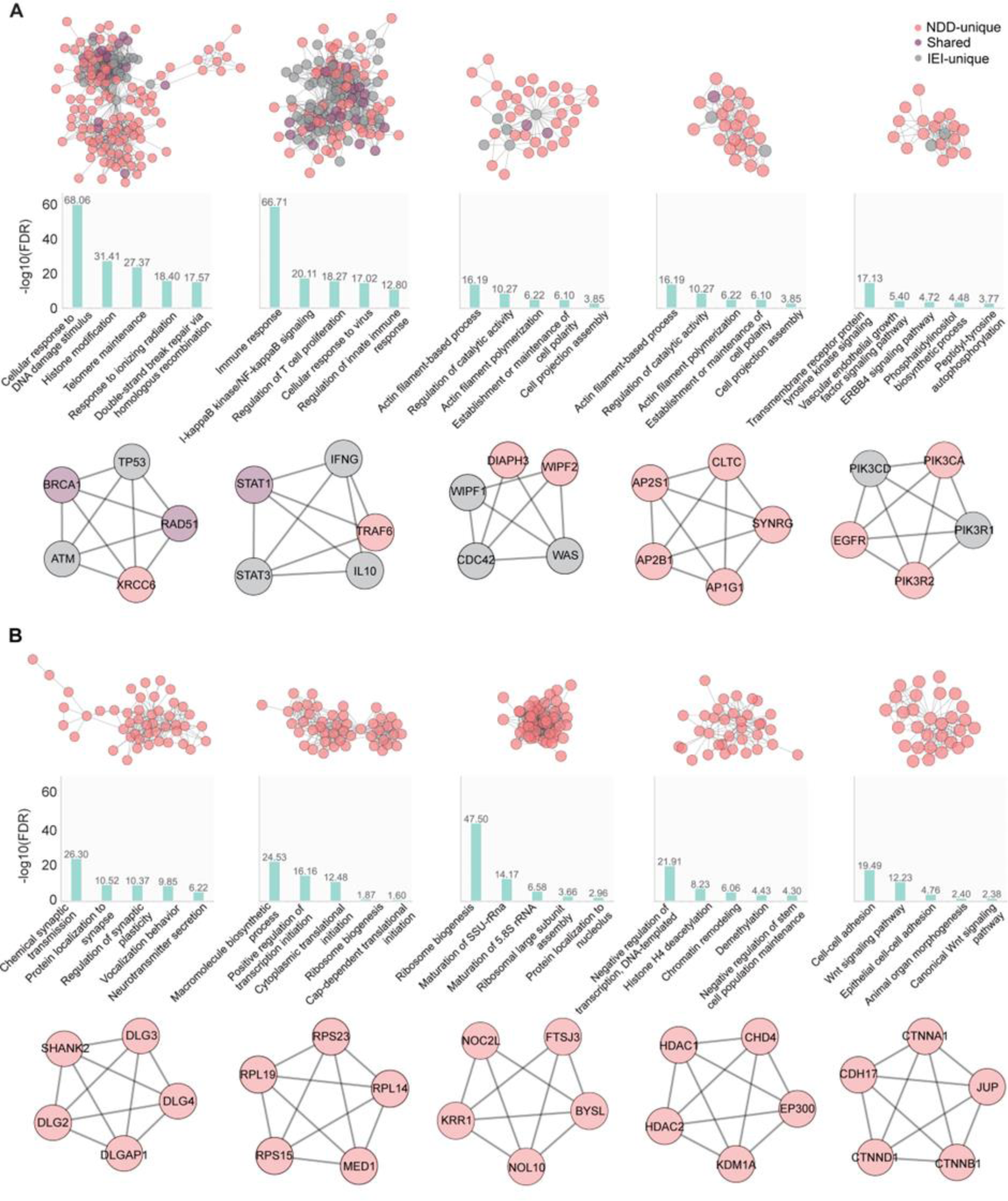
Protein network reconstruction integrates NDD-unique, IEI-unique and shared genes. **A)** Five major clusters with interactions between groups (top) with significantly enriched GO biological processes for each cluster (middle), and the five most central proteins within each cluster (bottom). **B)** Same as (A), but displaying clusters only for the NDD-unique gene products. Nodes represent proteins while edges indicate inferred interactions.

## Discussion

IEI are genetic disorders with variable phenotypic presentation. While IEIs are primarily characterized by the dysfunction or dysregulation of immune function, breakthroughs in patient treatment and care have led to better and longer life expectancy, enabling a more systematic identification of other phenotypes that persist even after immune system regularization. One of these are NDD-like symptoms, including social deficits, cognitive impairment and intellectual disability (Kurup et al. 2024).

While, in the past, these symptoms were often regarded as non-specific consequences of disease burden and repeated hospitalizations, an increasing number of studies have demonstrated that improving or even rescuing immune system function does not entirely improve these symptoms (Titman et al. 2008; Kerr et al. 2010; Nicholson et al. 2022). This suggests that NDDs might not just be a direct consequence of immune disease but could instead derive from specific effects that IEI-causing variants have in brain development and function. Here, we demonstrate that genes associated with both NDD and IEI phenotypes present with a number of molecular and network features that support an expression and functional overlap between immune and nervous systems. However, it remains a challenge, particularly within clinical populations, to undoubtedly differentiate between genetically-derived NDDs that arise during development and similar symptomatology secondary to an early IEI. Longitudinal measurements of neurodevelopmental milestones and cognitive function in IEI patients would be pivotal to answer this question, although the rarity of IEIs reduces the likelihood of capturing significant numbers of these patients in natural cohorts.

In this work, we used curated genotype-phenotype data to investigate whether genetic expression and function would support a duality of neuro-immune symptoms. We found that IEI-causing variants were frequently reported alongside NDD-like symptomatology, an overlap that ranged from 33% with the HGMD database, representing the full NDD spectrum, to 5.3% in the autism-specific variant database, SFARI. Notably, this symptom overlap cannot be directly translated into phenotype prevalence as we did not incorporate prevalence data in this study. Nonetheless, examining the genes involved in CVID phenotypes for instance (subtable 2; (Tangye et al. 2022)), representing the most common IEI (Ameratunga et al. 2023), we found 10 out of 23 causing genes in the HGMD database, suggesting that NDD symptoms are likely to be of clinical relevance to the IEI population. Furthermore, we have also identified IEI genes that have previously been associated with high NDD prevalence that were not yet described in the HGMD database. This is the case of *PIK3CD,* leading to APDS1, which is associated with an approximate 20% prevalence of NDD (Coulter et al. 2017; Serra et al. 2021; Oh et al. 2021). Altogether, our findings suggest that NDD symptomatology could be under-reported for a number of IEI-causing variants.

This interaction between immune system and brain function is not novel and has been repeatedly related with a number of disorders, including ASD and neurodegeneration (Q. Li and Barres 2017; Gupta et al. 2014). The most direct cellular link between these two systems is microglia, classically known as the resident immune cells of the brain. These cells, which maintain the immune homeostasis of the brain, are found in the foetal brain after migration from the yolk sac in the fourth postnatal week, and have been shown to proliferate in waves that closely follow neuronal proliferation patterns, facilitating typical brain development and neuronal wiring (Thion, Ginhoux, and Garel 2018; Paolicelli et al. 2011; A. R. Lawrence et al. 2024; Zhao, Umpierre, and Wu 2024; Menassa et al. 2022). Recent research however, puts into question the extent of the microglia relevance, as total absence of these cells from the neocortex and hippocampus resulted in typical development of brain function (O’Keeffe et al. 2025).

Dysfunctional activity of other immune cells, such as T cells, which can be found in meningeal tissue and the choroid plexus, could also contribute to altered brain development, particularly when cytokine release is involved (Schwartz et al. 2022). For instance, gestational TNF-ɑ and IL-1β levels, which modulate neuronal excitability, have been shown to later correlate with intellectual and cognitive scores, executive function and visuo-motor maturity in children, although the source of these cytokines is not yet fully ascertained (Zipp, Bittner, and Schafer 2023; Dozmorov et al. 2018; Ghassabian et al. 2018).

Importantly, many IEI-causing genes also have autonomous functions in the brain, independent of their roles within the immune system (Boulanger 2009). An example of this is the *PTEN* gene, which modulates neuronal size via the mTOR pathway and whose pathogenic variants frequently lead to cortical dysplasia and megalencephaly (Yehia, Keel, and Eng 2020; Shelkowitz et al. 2023). Another example are the DNA repair genes *NBN* and *ATM*, leading to Nijmegen Breakage Syndrome and Ataxia-Telangiectasia, respectively. Both of these genes are involved in brain developmental apoptosis, and abnormal activity of the mutated protein products appears to have a differential effect in distinct brain areas, particularly affecting the cerebellum (Rodrigues et al. 2013; Kose et al. 2024). Altogether, these diverse functions of IEI-causing genes indicate that compromised gene expression could, whether directly or indirectly, modulate brain developmental trajectories particularly during sensitive periods of development.

In order to provide early screening, it is imperative to identify which IEI patients would be at a higher risk of developing NDD phenotypes. Therapeutic interventions in NDDs are most successful when started early, when critical and sensitive periods of brain development are still open (J. Li et al. 2021). Behavioural therapy implementation at young ages was shown to have small to medium effects on reducing core NDD symptomatology (Shin, Park, and Lee 2025; Pe 2016). Furthermore, from preschool age onwards, formal behavioural and neurodevelopmental assessments can identify both clinical and sub-clinical cases in high-risk children who also present with high levels of mental health burden, alongside pre-existing NDD symptomatology (Boulton et al. 2023). These early assessments would be particularly relevant given that quality of life measures of children with NDDs appear to be primarily impacted by the presence of additional comorbidities (Mahjoob et al. 2024).

Here, we identified several genes that have been previously associated with both IEIs and NDDs and found that this combination of symptoms was enriched for genes involved in immune system development and DNA-repair pathways, suggesting that patients with variants along these pathways are more susceptible to the development of NDD symptomatology. Considering that genetic IEI and NDD databases are being continuously updated, future research could potentially identify other shared pathways between these patient populations. Although the association between these pathways and NDDs is not entirely novel, baseline psychiatry assessments are not common in IEIs, particularly when NDD manifestations are not evident. The genetic analysis performed here suggests that dedicated groups of IEI patients could benefit from a multidisciplinary team assessment at early diagnosis stages. Whilst a more systematic evaluation of NDDs in IEIs is required to ascertain the full spectrum and prevalence of NDDs in these patients, the inclusion of a neuropsychiatric evaluation as a standard assessment within distinct groups of IEIs could improve NDD diagnosis and decrease the disease burden that is associated with neurodevelopmental phenotypes.

## Methods

### Database mining

The last update of the Human Inborn Errors of Immunity (IEI) classification (Tangye et al. 2022), compiled by the International Union of Immunological Societies Expert Committee (IUIS), was used as the source of recognized causative IEI genes. All genetic entries, organized in 10 classification tables, were collected and then filtered for duplicates. In total, 449 unique genes were identified. To investigate the presence of NDD-like phenotypes reported in these tables, we considered entries where “*developmental delay*”, “*micro/macrocephaly*”, “*CNS abnormalities*”, “*cognitive and/or neurologic defects*”, “*mental/growth retardation*”, “*failure to thrive*”, and “*facial dysmorphism*” were indicated.

For the selection of neurodevelopmental disorder (NDD)-associated genes, the HGMD database (Stenson et al. 2020) was probed for “neurodevelopmental disorders”, originating a list of 18.507 disease-associated genes. After excluding duplicates and denominations not in compliance with the HGNC (Human Genome Organisation Gene Nomenclature Committee), a final list of 6003 NDD genes was obtained. Genes present in both gene lists were denominated as *shared*, while genes exclusive to the HGMD or IUIS lists were denominated as *NDD-unique* or *IEI-unique*, respectively.

For the investigation of additional NDD-associated gene databases, we resorted to the “Intellectual Disability and Autism Expert Panel” of ClinGen (clinicalgenome.org), which yielded 275 unique gene entries; the Geisinger Developmental Brain Disorder Gene Database that contemplates 732 curated genes involved in seven distinct disorders (*intellectual disability*, *autism*, *attention deficit hyperactivity disorder*, *schizophrenia*, *bipolar disorder*, *epilepsy*, and *cerebral palsy*) (Gonzalez-Mantilla et al. 2016); the Kyoto Encyclopedia of Genes and Genomes (KEGG; genome.jp/kegg) disease module for “neurodevelopmental disorder” which encompassed 354 unique gene entries; and the Simons Foundation Autism Research Initiative (SFARI; gene.sfari.org) gene database that contained 1162 autism spectrum disorder (ASD) risk genes.

To calculate the statistical significance of the overlap between distinct gene lists, the hypergeometric distribution probability was used(Bhandari et al. 2023). A possible total of 63.187 unique gene products was considered, following the last update of the GENCODE project (Frankish et al. 2021) (release 45 (GRCh38.p14), January 2024 update). This genome data includes protein-coding genes, long and small non-coding RNA genes, pseudogenes and immunoglobulin/T-cell receptor gene segments. To compare the number of observed shared genes and their expected frequency within the IEI classification tables, a Chi-squared test was performed. Graphs and data visualization were performed with GraphPad Prism 8 and InteractiVenn (Heberle et al. 2015).

### Gene ontology and functional enrichment analyses

Gene ontology (GO) analysis was used to investigate the enrichment of the identified genes of interest along distinct biological processes and pathways. GO analysis was performed using ShinyGO 0.80 (Ge, Jung, and Yao 2020), an online, open-access, platform that integrates classic GO functional categorizations with additional human pathway databases such as KEGG and the Molecular Signatures Database (MSigDB) (Liberzon et al. 2015). A False Discovery Rate (FDR) cutoff was defined at 0.05 and the minimal pathway size was defined at 10 to minimize term redundancy. The highest FDR calculated in the presented data was 1.60E-07. For visualization purposes, the top 10 significantly enriched terms were plotted as fold enrichment over the number of identified genes.

### Human RNAsequencing dataset preprocessing and analysis

All the 20.162 genes and expression profiles mapped by the Human Protein Atlas (HPA; proteinatlas.org) were downloaded and the genes of interest previously identified, belonging to the *NDD-unique*, *shared* and *IEI-unique* groups, were selected. When gene entries from the lists of interest did not match HPA database entries, gene names were manually checked for possible synonym names within the HGNC database. The majority of the non-identified genes encoded for non-coding RNA genes or pseudogenes. In total, we selected RNA sequencing data (bulk and single-cell) for 5798 *NDD-unique*, 150 *shared* and 264 *IEI-unique* protein coding genes, including their expression profiles across 50 tissue types, 13 brain areas, 19 blood cell types and 81 cell types. For further analysis, expression values under 30 normalized transcripts per million (nTPM) were removed and considered as noise, while the remaining values were log normalized. We quantified gene products enriched on tissues of interest using the RNA tissue-specific data as in (Digre and Lindskog 2021).

Comparative gene expression between groups was analyzed, for each comparison pair log2 fold differences were calculated from mean protein abundances and statistical significance was assessed using independent t-tests. P-values were adjusted for multiple comparisons using the Benjamini-Hochberg false discovery rate (FDR) correction implemented in statsmodels (v0.14.1) through python (v3.12.8). Volcano plots were generated with log2 fold differences on the x-axis and -log10(FDR) on the y-axis, with most significant genes annotated (FDR < 0.05). P-values for all paired comparisons are reported in Supplementary tables 2-5.

### Protein interaction clustering analysis

Protein-protein interaction networks were constructed and analyzed using Cytoscape (v3.10.2) (Shannon et al. 2003). High-confidence protein interactions (confidence score ≥ 0.7) were retrieved from the STRING database (v12.0) for all three gene groups (Szklarczyk et al. 2023).

To identify functionally related protein clusters, we employed the Markov Cluster Algorithm with a granularity parameter of 4, which allowed for the detection of highly interconnected regions likely corresponding to shared functional domains.

The biological significance of identified protein-protein interaction clusters was assessed through enrichment analysis using the STRING enrichment tool. The five largest intersecting clusters with at least two connections between protein groups were selected. From these clusters, we extracted the enriched GO biological processes and removed similar terms using the Hobohm method using a redundancy cutoff of 0.3 (Hobohm et al. 1992).

We reported the top 5 enriched GO biological processes per cluster using a FDR-corrected p-value < 0.05. The enrichment analysis accounted for the complete set of proteins in the STRING database as the background reference set. The pathway enrichment results were visualized using custom Python (v3.12.8) scripts. Central nodes in each cluster were determined using Maximal Clique Centrality incorporated into cytoHubba (v0.1) (Chin et al. 2014).

### Gene association visualization and classification

Gene expression data was obtained from the HPA Cell Atlas in log-transformed format. This was then complemented with the previously defined gene classifications: *IEI-unique* or *shared* genes, in addition to either *NDD-unique* genes (from the HGMD database) or autism-risk genes exclusively categorized as syndromic within the SFARI database (*SFARI-S*).

We performed dimensionality reduction using Uniform Manifold Approximation and Projection (UMAP) (McInnes et al., 2018) to visualize high-dimensional gene expression patterns. The UMAP algorithm was configured with cosine distance as the metric and the following parameters: n_neighbors = 20, min_dist = 0.1, and n_components = 2. Two-dimensional embeddings were generated and visualized using matplotlib.

For supervised classification analysis, we extracted gene expression features while excluding categorical variables. Genes were labeled as *IEI-unique*, *shared*, or *SFARI-S*. Using stratified sampling to preserve class proportions, we split the dataset into training (80%) and testing (20%) sets. We applied a standard scaler transformation to normalize feature distributions. A Random Forest Classifier comprising 100 decision trees was trained on the standardized data to predict gene group membership in the test set. Model performance evaluation included the multiple classification metrics precision, recall, and F1-score. To assess classification performance for each gene group, we implemented a one-vs-rest multiclass analysis and generated receiver operating characteristic (ROC) curves and calculated corresponding area under the curve (AUC) values.

## Data Availability

All data used in our paper are already public and can be downloaded from their respective repositories (see Methods). The analysis code is openly available at https://github.com/BaduraLab/GeneticOverlap-IEI-NDD

## Acknowledgements

This research was funded by the Erasmus MC Starting Grant (IS) and the Convergence Health & Technology Integrative Neuromedicine Flagship grant (AB). PJS has received funding from the European Union’s Horizon 2020 research and innovation programme under grant agreement No 779295.for the ImmunAID project. VASHD received funding from ZonMw, Horizon 2020, Pharming, CSL Behring : Research; Takeda, CSL Behring, Jansen & Jansen NL, Astra Zeneca, Moderna : speaker’s fee / honoraria.

