## Supplementary figures and images for "Analysis of genetic overlap between inborn errors of immunity and neurodevelopmental disorders"

### Supplementary Figure 1

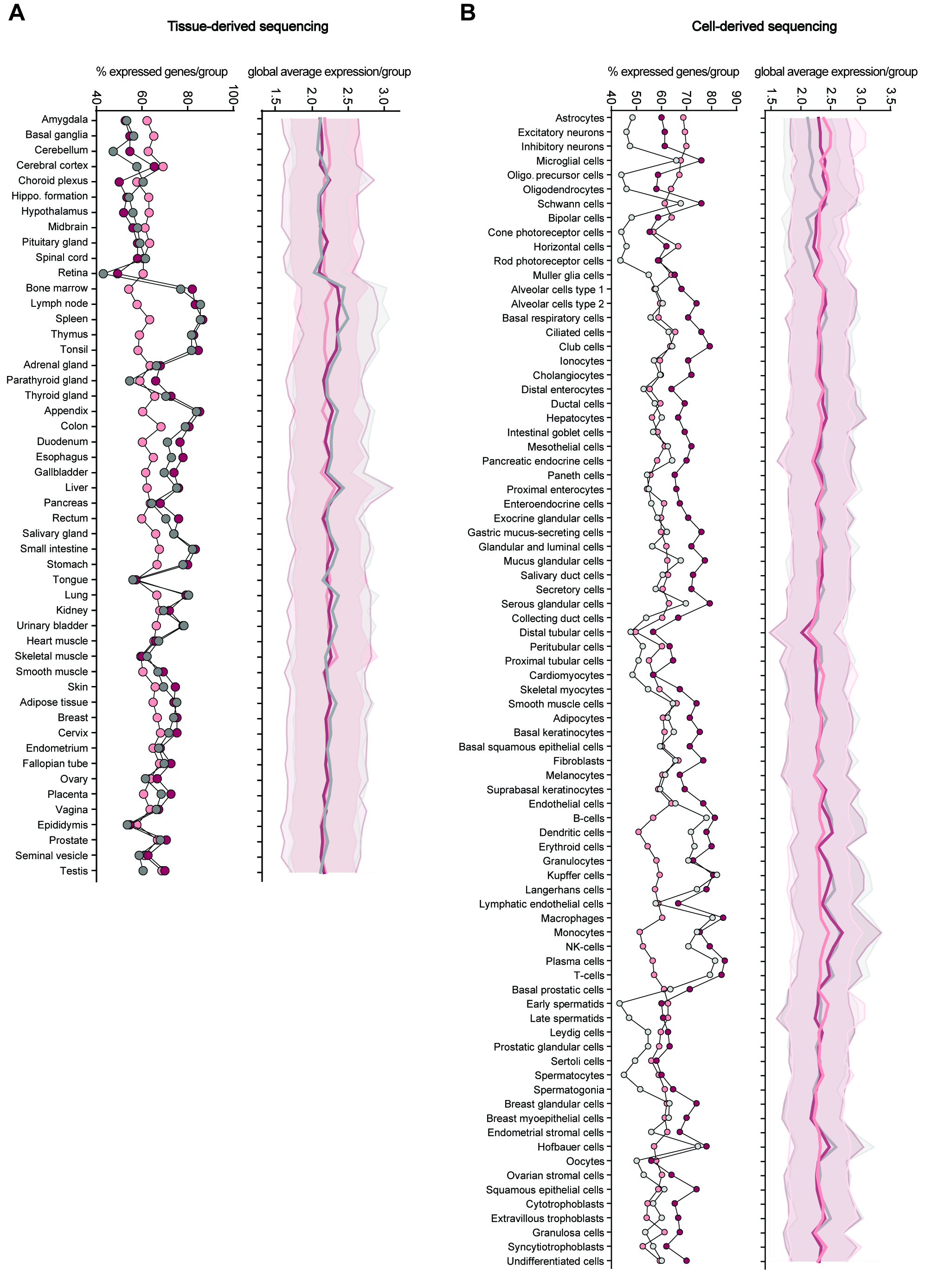

### Supplementary Figure 2

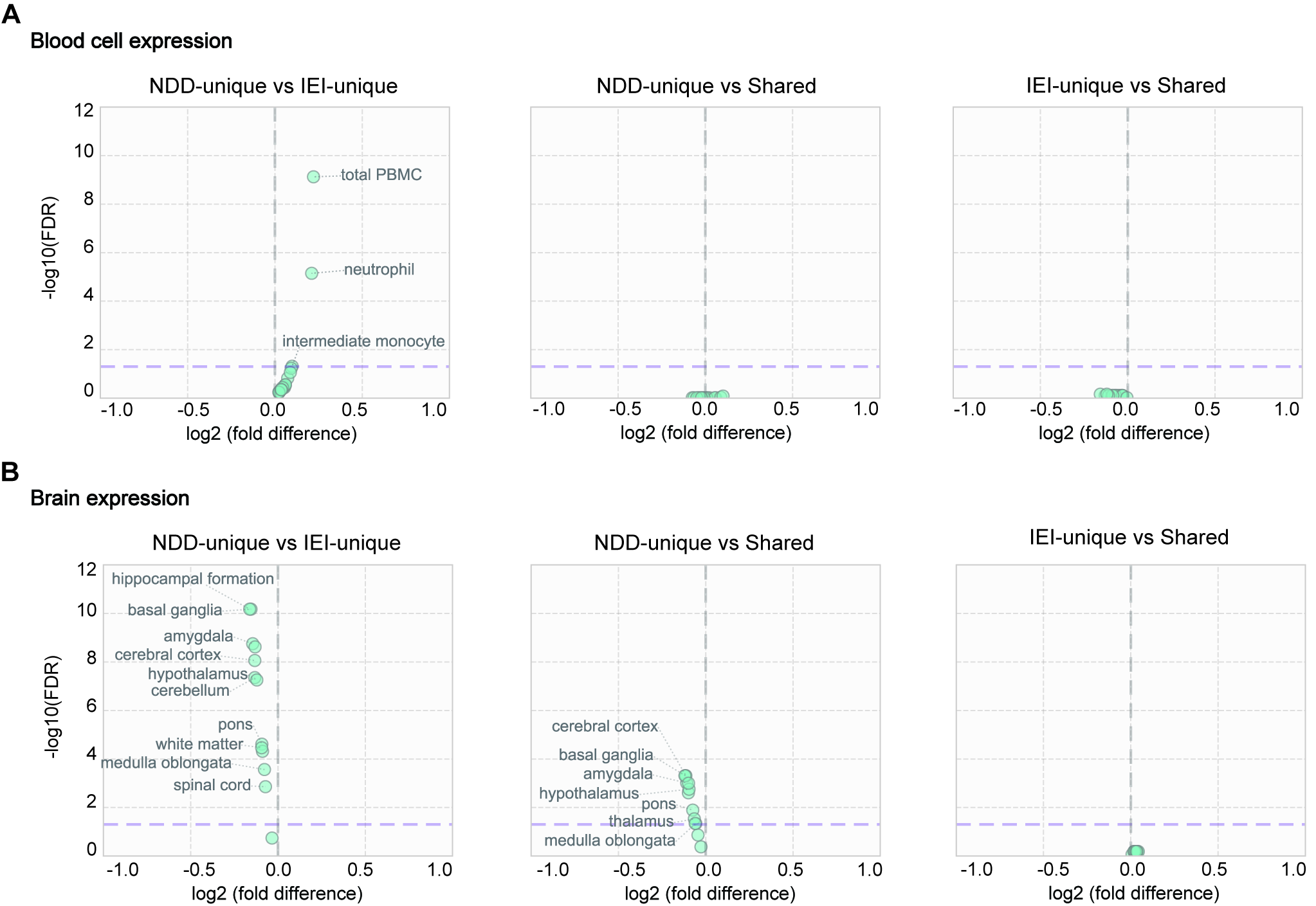
