## Supplementary Tables for "Analysis of genetic overlap between inborn errors of immunity and neurodevelopmental disorders"

| **Table 1** | Immunodeficiencies affecting cellular and humoral immunity |
| --- | --- |
| **Table 2** | Combined immunodeficiencies with associated or syndromic features |
| **Table 3** | Predominantly Antibody Deficiencies |
| **Table 4** | Diseases of Immune Dysregulation |
| **Table 5** | Congenital defects of phagocyte number or function |
| **Table 6** | Defects in Intrinsic and Immunity |
| **Table 7** | Autoinflammatory Disorders |
| **Table 8** | Complement Deficiencies |
| **Table 9** | Bone Marrow Failure |
| **Table 10** | Phenocopies of Inborn Errors of Immunity |

**Supplementary Table 1:  *IEI classification.*** IEI grouping employed by the International Union of Immunological Societies (IUIS) Expert Committee.

| **Single-cell dataset** | | | | | | |
| --- | --- | --- | --- | --- | --- | --- |
| **Feature** | **NDD-unique vs IEI-unique** | | **NDD-unique vs shared** | | **IEI-unique vs shared** | |
|  | log2 fold diff. | FDR | log2 fold diff. | FDR | log2 fold diff. | FDR |
| Adipocytes | 0,125 | 0,013 | 0,138 | 0,049 | 0,013 | 0,910 |
| Alveolar cells type 1 | 0,028 | 0,699 | 0,131 | 0,089 | 0,103 | 0,337 |
| Alveolar cells type 2 | 0,065 | 0,287 | 0,212 | 0,003 | 0,147 | 0,176 |
| Astrocytes | -0,516 | 2,13E-06 | -0,237 | 0,001 | 0,279 | 0,038 |
| B-cells | 0,421 | 3,95E-06 | 0,397 | 2,53E+06 | -0,024 | 0,746 |
| Basal keratinocytes | 0,111 | 0,023 | 0,176 | 0,007 | 0,065 | 0,423 |
| Basal prostatic cells | 0,073 | 0,181 | 0,139 | 0,041 | 0,067 | 0,431 |
| Basal respiratory cells | 0,006 | 0,921 | 0,136 | 0,071 | 0,130 | 0,322 |
| Basal squamous epithelial cells | 0,007 | 0,917 | 0,105 | 0,139 | 0,097 | 0,323 |
| Bipolar cells | -0,385 | 1,94E+02 | -0,188 | 0,009 | 0,197 | 0,177 |
| Breast glandular cells | 0,029 | 0,618 | 0,088 | 0,174 | 0,059 | 0,489 |
| Breast myoepithelial cells | 0,035 | 0,551 | 0,071 | 0,276 | 0,036 | 0,687 |
| Cardiomyocytes | -0,171 | 0,007 | -0,046 | 0,576 | 0,124 | 0,323 |
| Cholangiocytes | 0,060 | 0,402 | 0,207 | 0,009 | 0,147 | 0,267 |
| Ciliated cells | 9,28E+10 | 0,998 | 0,095 | 0,105 | 0,094 | 0,320 |
| Club cells | 0,071 | 0,164 | 0,166 | 0,007 | 0,095 | 0,322 |
| Collecting duct cells | -0,084 | 0,218 | 0,068 | 0,401 | 0,152 | 0,320 |
| Cone photoreceptor cells | -0,448 | 3,86E+07 | -0,150 | 0,135 | 0,298 | 0,133 |
| Cytotrophoblasts | 0,055 | 0,472 | 0,086 | 0,319 | 0,031 | 0,825 |
| Dendritic cells | 0,477 | 0,006 | 0,484 | 7,94E+05 | 0,008 | 0,938 |
| Distal enterocytes | -0,022 | 0,800 | 0,111 | 0,193 | 0,133 | 0,323 |
| Distal tubular cells | -0,057 | 0,633 | 0,041 | 0,747 | 0,098 | 0,627 |
| Ductal cells | 0,008 | 0,917 | 0,147 | 0,049 | 0,140 | 0,320 |
| Early spermatids | -0,359 | 3,72E+04 | -0,169 | 0,025 | 0,190 | 0,177 |
| Endometrial stromal cells | -0,069 | 0,218 | 0,067 | 0,319 | 0,136 | 0,290 |
| Endothelial cells | 0,132 | 0,001 | 0,148 | 0,009 | 0,016 | 0,850 |
| Enteroendocrine cells | -0,087 | 0,164 | 0,026 | 0,724 | 0,113 | 0,323 |
| Erythroid cells | 0,364 | 2,47E+04 | 0,399 | 1,16E+09 | 0,035 | 0,655 |
| Excitatory neurons | -0,615 | 1,67E-12 | -0,244 | 0,001 | 0,371 | 0,005 |
| Exocrine glandular cells | 0,012 | 0,894 | 0,138 | 0,058 | 0,126 | 0,322 |
| Extravillous trophoblasts | 0,175 | 0,004 | 0,154 | 0,069 | -0,021 | 0,886 |
| Fibroblasts | 0,072 | 0,096 | 0,099 | 0,071 | 0,028 | 0,727 |
| Gastric mucus-secreting cells | 0,065 | 0,280 | 0,214 | 0,002 | 0,149 | 0,133 |
| Glandular and luminal cells | -0,031 | 0,618 | 0,096 | 0,150 | 0,128 | 0,302 |
| Granulocytes | 0,258 | 6,10E+07 | 0,269 | 6,60E+10 | 0,011 | 0,917 |
| Granulosa cells | -0,161 | 0,002 | 0,003 | 0,962 | 0,163 | 0,210 |
| Hepatocytes | 0,168 | 0,004 | 0,227 | 0,004 | 0,059 | 0,585 |
| Hofbauer cells | 0,386 | 4,21E-01 | 0,334 | 8,59E+08 | -0,051 | 0,585 |
| Horizontal cells | -0,485 | 2,11E-03 | -0,233 | 0,001 | 0,252 | 0,117 |
| Inhibitory neurons | -0,593 | 2,12E-12 | -0,248 | 0,001 | 0,345 | 0,005 |
| Intestinal goblet cells | -0,006 | 0,921 | 0,123 | 0,113 | 0,129 | 0,322 |
| Ionocytes | -0,028 | 0,687 | 0,124 | 0,089 | 0,152 | 0,287 |
| Kupffer cells | 0,440 | 1,61E-07 | 0,388 | 4,58E+06 | -0,052 | 0,431 |
| Langerhans cells | 0,365 | 3,16E-01 | 0,307 | 1,73E+10 | -0,058 | 0,483 |
| Late spermatids | -0,278 | 7,14E+07 | -0,153 | 0,038 | 0,125 | 0,323 |
| Leydig cells | -0,080 | 0,165 | 0,038 | 0,581 | 0,118 | 0,323 |
| Lymphatic endothelial cells | 0,045 | 0,534 | 0,105 | 0,176 | 0,060 | 0,603 |
| Macrophages | 0,360 | 3,90E-03 | 0,339 | 1,37E+07 | -0,021 | 0,799 |
| Melanocytes | 0,040 | 0,594 | 0,071 | 0,391 | 0,031 | 0,825 |
| Mesothelial cells | 0,098 | 0,056 | 0,156 | 0,021 | 0,058 | 0,551 |
| Microglial cells | -0,117 | 0,015 | 0,104 | 0,089 | 0,221 | 0,020 |
| Monocytes | 0,482 | 0,007 | 0,479 | 7,37E+05 | -0,003 | 0,978 |
| Mucus glandular cells | 0,081 | 0,114 | 0,194 | 0,002 | 0,114 | 0,187 |
| Muller glia cells | -0,144 | 0,003 | -0,043 | 0,520 | 0,101 | 0,323 |
| NK-cells | 0,450 | 0,420 | 0,498 | 8,52E+04 | 0,048 | 0,603 |
| Oligodendrocyte precursor cells | -0,570 | 9,07E-09 | -0,197 | 0,007 | 0,373 | 0,005 |
| Oligodendrocytes | -0,498 | 0,005 | -0,173 | 0,025 | 0,325 | 0,020 |
| Oocytes | -0,204 | 0,001 | -0,066 | 0,402 | 0,138 | 0,323 |
| Ovarian stromal cells | -0,083 | 0,165 | 0,011 | 0,870 | 0,094 | 0,423 |
| Pancreatic endocrine cells | 0,155 | 0,009 | 0,171 | 0,039 | 0,016 | 0,910 |
| Paneth cells | 0,024 | 0,772 | 0,163 | 0,055 | 0,139 | 0,322 |
| Peritubular cells | -0,147 | 0,010 | -0,067 | 0,387 | 0,080 | 0,569 |
| Plasma cells | 0,403 | 4,24E-04 | 0,404 | 8,52E+04 | 0,001 | 0,980 |
| Prostatic glandular cells | -0,059 | 0,372 | 0,046 | 0,543 | 0,105 | 0,323 |
| Proximal enterocytes | 0,013 | 0,913 | 0,151 | 0,089 | 0,138 | 0,323 |
| Proximal tubular cells | 0,010 | 0,917 | 0,103 | 0,206 | 0,093 | 0,424 |
| Rod photoreceptor cells | -0,414 | 2,00E+04 | -0,156 | 0,054 | 0,258 | 0,118 |
| Salivary duct cells | 0,007 | 0,917 | 0,152 | 0,021 | 0,145 | 0,133 |
| Schwann cells | 0,184 | 0,000 | 0,231 | 0,001 | 0,047 | 0,603 |
| Secretory cells | 0,037 | 0,554 | 0,125 | 0,069 | 0,088 | 0,364 |
| Serous glandular cells | 0,121 | 0,011 | 0,197 | 0,002 | 0,076 | 0,323 |
| Sertoli cells | -0,162 | 0,015 | -0,020 | 0,811 | 0,142 | 0,323 |
| Skeletal myocytes | -0,048 | 0,485 | 0,077 | 0,311 | 0,125 | 0,295 |
| Smooth muscle cells | 0,038 | 0,452 | 0,071 | 0,206 | 0,033 | 0,665 |
| Spermatocytes | -0,303 | 6,10E+07 | -0,046 | 0,543 | 0,257 | 0,058 |
| Spermatogonia | -0,196 | 1,66E+11 | -0,024 | 0,701 | 0,172 | 0,153 |
| Squamous epithelial cells | 0,034 | 0,618 | 0,135 | 0,076 | 0,100 | 0,323 |
| Suprabasal keratinocytes | 0,052 | 0,415 | 0,151 | 0,040 | 0,099 | 0,323 |
| Syncytiotrophoblasts | 0,080 | 0,264 | 0,066 | 0,455 | -0,014 | 0,922 |
| T-cells | 0,384 | 0,001 | 0,379 | 1,59E+05 | -0,005 | 0,938 |
| Undifferentiated cells | -0,025 | 0,726 | 0,111 | 0,135 | 0,136 | 0,320 |

**Supplementary Table 2: *Single-cell dataset analysis.*** Log2 fold difference and false discovery rate values for all paired comparisons between NDD-unique, shared and IEI-unique gene expression values within the single-cell HPA dataset.

| Tissue dataset | | | | | | |
| --- | --- | --- | --- | --- | --- | --- |
| **Feature** | **NDD-unique vs IEI-unique** | | **NDD-unique vs Shared** | | **IEI-unique vs Shared** | |
|  | log2 fold diff. | FDR | log2 fold diff. | FDR | log2 fold diff. | FDR |
| Adipose tissue | 0,099 | 0,007 | 0,069 | 0,260 | -0,030 | 0,992 |
| Adrenal gland | 0,006 | 0,903 | 0,026 | 0,709 | 0,020 | 0,992 |
| Amygdala | -0,215 | 3,42E+08 | -0,209 | 0,001 | 0,005 | 0,992 |
| Appendix | 0,272 | 15,035 | 0,268 | 4,30E+07 | -0,004 | 0,992 |
| Basal ganglia | -0,237 | 8,80E+06 | -0,211 | 0,001 | 0,026 | 0,992 |
| Bone marrow | 0,333 | 1,48E+04 | 0,312 | 6,18E+09 | -0,020 | 0,992 |
| Breast | 0,029 | 0,432 | 0,030 | 0,652 | 0,001 | 0,997 |
| Cerebellum | -0,392 | 0,133 | -0,244 | 0,000 | 0,148 | 0,992 |
| Cerebral cortex | -0,218 | 8,80E+06 | -0,156 | 0,005 | 0,062 | 0,992 |
| Cervix | -0,005 | 0,903 | 0,025 | 0,709 | 0,031 | 0,992 |
| Choroid plexus | -0,024 | 0,617 | -0,073 | 0,322 | -0,049 | 0,992 |
| Colon | 0,057 | 0,097 | 0,057 | 0,294 | 0,000 | 0,997 |
| Duodenum | 0,126 | 0,001 | 0,151 | 0,006 | 0,025 | 0,992 |
| Endometrium | -0,027 | 0,511 | 0,013 | 0,820 | 0,039 | 0,992 |
| Epididymis | -0,099 | 0,026 | -0,056 | 0,474 | 0,043 | 0,992 |
| Esophagus | 0,041 | 0,302 | 0,087 | 0,124 | 0,046 | 0,992 |
| Fallopian tube | 0,004 | 0,910 | 0,037 | 0,591 | 0,032 | 0,992 |
| Gallbladder | 0,084 | 0,024 | 0,139 | 0,007 | 0,055 | 0,992 |
| Heart muscle | -0,037 | 0,390 | -0,043 | 0,587 | -0,005 | 0,992 |
| Hippocampal formation | -0,223 | 8,78E+07 | -0,204 | 0,001 | 0,019 | 0,992 |
| Hypothalamus | -0,208 | 3,21E+09 | -0,195 | 0,001 | 0,013 | 0,992 |
| Kidney | -0,009 | 0,842 | 0,024 | 0,709 | 0,033 | 0,992 |
| Liver | 0,168 | 1,67E+10 | 0,176 | 0,002 | 0,008 | 0,992 |
| Lung | 0,174 | 1,62E+08 | 0,143 | 0,004 | -0,030 | 0,992 |
| Lymph node | 0,342 | 2,74E-04 | 0,338 | 6,88E+05 | -0,004 | 0,992 |
| Midbrain | -0,157 | 0,000 | -0,153 | 0,011 | 0,004 | 0,992 |
| Ovary | -0,064 | 0,136 | -0,014 | 0,820 | 0,050 | 0,992 |
| Pancreas | -0,044 | 0,299 | 0,031 | 0,657 | 0,075 | 0,992 |
| Parathyroid gland | -0,124 | 0,011 | 0,020 | 0,794 | 0,144 | 0,862 |
| Pituitary gland | -0,153 | 5,78E+11 | -0,060 | 0,332 | 0,093 | 0,992 |
| Placenta | 0,069 | 0,111 | 0,093 | 0,158 | 0,023 | 0,992 |
| Prostate | -0,031 | 0,398 | -0,020 | 0,743 | 0,011 | 0,992 |
| Rectum | 0,089 | 0,026 | 0,144 | 0,009 | 0,055 | 0,992 |
| Retina | -0,450 | 0,000 | -0,248 | 0,000 | 0,202 | 0,776 |
| Salivary gland | 0,040 | 0,299 | 0,067 | 0,251 | 0,026 | 0,992 |
| Seminal vesicle | -0,078 | 0,068 | -0,007 | 0,886 | 0,071 | 0,992 |
| Skeletal muscle | -0,118 | 0,013 | -0,087 | 0,265 | 0,031 | 0,992 |
| Skin | -0,027 | 0,494 | 0,032 | 0,651 | 0,059 | 0,992 |
| Small intestine | 0,160 | 2,67E+07 | 0,149 | 0,001 | -0,012 | 0,992 |
| Smooth muscle | 0,038 | 0,390 | 0,072 | 0,275 | 0,034 | 0,992 |
| Spinal cord | -0,099 | 0,019 | -0,093 | 0,158 | 0,006 | 0,992 |
| Spleen | 0,331 | 0,000 | 0,251 | 8,89E+08 | -0,080 | 0,992 |
| Stomach | 0,083 | 0,013 | 0,094 | 0,051 | 0,012 | 0,992 |
| Testis | -0,153 | 1,47E+11 | -0,009 | 0,850 | 0,144 | 0,616 |
| Thymus | 0,275 | 3,95E+03 | 0,310 | 7,18E+06 | 0,035 | 0,992 |
| Thyroid gland | -0,001 | 0,986 | 0,033 | 0,651 | 0,034 | 0,992 |
| Tongue | -0,135 | 0,008 | -0,028 | 0,743 | 0,107 | 0,992 |
| Tonsil | 0,304 | 15,035 | 0,338 | 6,88E+05 | 0,034 | 0,992 |
| Urinary bladder | 0,129 | 0,000 | 0,110 | 0,035 | -0,020 | 0,992 |
| Vagina | -0,039 | 0,361 | 0,017 | 0,794 | 0,056 | 0,992 |

**Supplementary Table 3: *Tissue dataset analysis.*** Log2 fold difference and false discovery rate values for all paired comparisons between NDD-unique, shared and IEI-unique gene expression valued within the tissue HPA dataset.

| Blood dataset | | | | | | |
| --- | --- | --- | --- | --- | --- | --- |
| **Feature** | **NDD-unique vs IEI-unique** | | **NDD-unique vs shared** | | **IEI-unique vs shared** | |
|  | log2 fold diff. | FDR | log2 fold diff. | FDR | log2 fold diff. | FDR |
| Basophil | 0,055 | 0,367 | -0,075 | 0,959 | -0,129 | 0,776 |
| Classical monocyte | 0,073 | 0,151 | 0,024 | 0,959 | -0,049 | 0,777 |
| Eosinophil | 0,024 | 0,566 | -0,042 | 0,959 | -0,066 | 0,776 |
| GdT-cell | 0,054 | 0,325 | -0,017 | 0,959 | -0,070 | 0,776 |
| Intermediate monocyte | 0,099 | 0,048 | 0,059 | 0,959 | -0,039 | 0,777 |
| MAIT T-cell | 0,035 | 0,446 | 0,002 | 0,976 | -0,033 | 0,777 |
| Memory B-cell | 0,048 | 0,367 | -0,010 | 0,959 | -0,059 | 0,776 |
| Memory CD4 T-cell | 0,059 | 0,281 | -0,027 | 0,959 | -0,086 | 0,776 |
| Memory CD8 T-cell | 0,060 | 0,281 | -0,016 | 0,959 | -0,076 | 0,776 |
| Myeloid DC | 0,043 | 0,367 | 0,010 | 0,959 | -0,034 | 0,777 |
| Naive B-cell | 0,036 | 0,446 | 0,007 | 0,959 | -0,029 | 0,777 |
| Naive CD4 T-cell | 0,025 | 0,566 | -0,065 | 0,959 | -0,090 | 0,776 |
| Naive CD8 T-cell | 0,036 | 0,446 | -0,035 | 0,959 | -0,071 | 0,776 |
| Neutrophil | 0,211 | 7,11E+09 | 0,053 | 0,959 | -0,157 | 0,685 |
| NK-cell | 0,022 | 0,566 | -0,008 | 0,959 | -0,030 | 0,777 |
| Non-classical monocyte | 0,093 | 0,059 | 0,089 | 0,915 | -0,005 | 0,952 |
| Plasmacytoid DC | 0,036 | 0,446 | -0,049 | 0,959 | -0,085 | 0,776 |
| T-reg | 0,088 | 0,087 | -0,022 | 0,959 | -0,110 | 0,776 |
| Total PBMC | 0,221 | 7,48E+05 | 0,101 | 0,819 | -0,120 | 0,685 |

**Supplementary Table 4: *Blood dataset analysis.*** Log2 fold difference and false discovery rate values for all paired comparisons between NDD-unique, shared and IEI-unique gene expression values within the blood HPA dataset.

| Brain dataset | | | | | | |
| --- | --- | --- | --- | --- | --- | --- |
| **Feature** | **NDD-unique vs IEI-unique** | | **NDD-unique vs shared** | | **IEI-unique vs shared** | |
|  | log2 fold diff. | FDR | log2 fold diff. | FDR | log2 fold diff. | FDR |
| Amygdala | -0,145 | 1,77E+07 | -0,109 | 0,001 | 0,036 | 0,652 |
| Basal ganglia | -0,155 | 6,65E+04 | -0,117 | 0,000 | 0,038 | 0,652 |
| Cerebellum | -0,133 | 4,42E+07 | -0,099 | 0,002 | 0,034 | 0,652 |
| Cerebral cortex | -0,133 | 8,68E+05 | -0,114 | 0,000 | 0,019 | 0,652 |
| Choroid plexus | -0,034 | 0,181 | -0,027 | 0,418 | 0,007 | 0,865 |
| Hippocampal formation | -0,162 | 6,65E+04 | -0,119 | 0,000 | 0,043 | 0,652 |
| Hypothalamus | -0,121 | 5,53E+08 | -0,095 | 0,002 | 0,026 | 0,652 |
| Medulla oblongata | -0,078 | 0,000 | -0,057 | 0,046 | 0,021 | 0,652 |
| Midbrain | -0,131 | 2,39E+06 | -0,098 | 0,001 | 0,033 | 0,652 |
| Pons | -0,093 | 2,45E+11 | -0,074 | 0,013 | 0,018 | 0,652 |
| Spinal cord | -0,072 | 0,001 | -0,045 | 0,136 | 0,027 | 0,652 |
| Thalamus | -0,089 | 4,81E+10 | -0,065 | 0,029 | 0,024 | 0,652 |
| White matter | -0,094 | 3,43E+11 | -0,060 | 0,046 | 0,033 | 0,652 |

**Supplementary Table 5: *Brain dataset analysis.*** Log2 fold difference and false discovery rate values for all paired comparisons between NDD-unique, shared and IEI-unique gene expression values within the brain HPA dataset.
